# INVESTIGATING RATIONAL ANTIBIOTIC PRESCRIPTIONS IN PUBLIC HEALTH CARE FACILITIES OF ZANZIBAR,TANZANIA

**DOI:** 10.64898/2026.09.09.26362003

**Authors:** Mayasa Salum, Omary Swallehe

## Abstract

This study aims at assessing the rational antibiotic medicines prescribing in public health care facilities in Zanzibar, Tanzania. The study employed a descriptive and cross-sectional study with a quantitative approach in collecting and analysing data. The study targeted all 30 health care facilities in the western urban region of Zanzibar together with all prescribers and prescriptions prescribed during the study period. Prescriptions were selected by using probability systematic sampling techniques from selected public health facilities.

The study found that, 3,455 (99.9%) recorded prescription encounters had a documented diagnosis, indicating comprehensive record-keeping. On average, 1.1 (SD= ±0.3) antibiotics were prescribed per prescription encounter, with 2,277 (65.9%) prescribed generic names. Injectable antibiotics were recorded in 166 (4.8%) of prescriptions, while 2,270 (65.7%) of prescriptions adhered to the Zanzibar Standard Treatment Guidelines (STG). The overall mean knowledge score among prescribers was 6.1 (SD = 0.89). 24.3% of prescribers had low knowledge (≤5 points), 41.4%had medium knowledge (6 points), and 34.3%had high knowledge (≥7 points). A majority of prescribers (90%) reported that they always adhere to the Zanzibar Standard Treatment Guidelines (ZSTG) and Zanzibar Essential Medicines List (ZEML) when prescribing antibiotics. 1.4% of prescribers acknowledged that they sometimes prescribe antibiotics even when unnecessary. Regarding antibiotic selection, 45.7% of prescribers prescribe only generic antibiotics. The study also found that Beta-Lactam antibiotics were the most prescribed, with 98.6% of prescribers confirming their frequent use. According to the results, most of the prescribing indicators in these health facilities did not meet the WHO’s standard requirements, thus irrational (poor rational prescribing).

## 1. Introduction

The rational use of antibiotics requires the selection of an appropriate antibiotic and its administration by the most effective route at the correct dose, frequency, and duration. (1) Several factors, such as years of experience, educational background, and training in rational medicine use, influence health professionals’ prescription of rational medicines. (2) According to WHO estimates, half of patients do not take their medications as recommended, and over half are improperly sold, administered, or prescribed. (3) A set of medicine use indicators recommended by the WHO has been shown to help examine the practices of prescribing medicines in healthcare facilities. Many studies that have examined issues with medicine prescribing patterns across the globe have included the use of prescribing indicators; WHO indicators include the average number of drugs prescribed for each encounter, the proportion of prescriptions written under generic names, the percentage of prescriptions for antibiotics, the percentage of prescriptions for injections, and the percentage of prescriptions from the EML.(2) Nowadays, antibiotics are the most often given medications in hospitals worldwide, and their role in treating infections cannot be overstated. However, inappropriate antibiotic use leads to bacterial resistance, significantly influencing treatment success and speeding up the formation and spread of resistant microorganisms.(3) Because antibiotics are more likely to promote Antimicrobial Resistance (AMR), they should be prescribed, dispensed, and administered with greater caution than other medications. (4) When antibiotics are used wisely, healthcare expenses can be decreased, as can the possibility of side effects for the patient. The broad environmental influences that cause bacteria to become antibiotic-resistant can also be reduced. (2) Irrational antibiotic usage is a problem, particularly in underdeveloped countries. Medicine prescribing, dispensing, patient compliance, and irrational prescriptions and medicine use are common in healthcare settings of developing countries. These include the use of multiple prescription medications, prescribing medications against clinical guidelines, allowing patients to self-medicate, overusing antibiotics and injectables, and using medicines with questionable origins.

Guidelines on the use of Antibiotics are essential to consider when prescribing antibiotics. The doctor/prescriber should examine the possibility of antibiotic side effects/ adverse reactions as well as potential drug-drug or drug-food interactions. Antibiotics are categorized into three groups in the AWaRe tool: Access, and Watch, Reserve. Based on each category’s specific impact on AMR. Because of their limited range of action, “Access” antibiotics usually have fewer side effects, a lesser chance of antimicrobial resistance developing, and are less expensive. On the other hand, “Watch” antibiotics are usually administered for patients with more serious diseases, mostly in hospital settings. They have a higher risk of fostering antimicrobial resistance. However, “Reserve” antibiotics are regarded as a last choice and ought to be used only in cases of severe illnesses brought on by microorganisms resistant to several drugs. Globally, the prevalence of rational use of antibiotic prescribing was 74.6% in Canada and 88% in London. (5) (6). The prevalence of rational use of antibiotic prescribing across sub-Saharan Africa was 75.6% in Nigeria, 53% in Eritrea and 69.7% in Ethiopia. (2) (4) (7), and in National, the prevalence of rational use of antibiotic prescribing was 74.4% in Tanzania.

The scope of this study (assessing the rational use of antibiotic prescribing) typically includes evaluating adherence to clinical guidelines, appropriateness of antibiotic selection, dosage, duration, and frequency, and the effects of using antibiotics on patient outcomes, antimicrobial resistance patterns, and healthcare costs. This study examines prescribing practices across various healthcare settings and identifies areas for improvement to promote optimal antibiotic use and combat antimicrobial resistance.

Antimicrobial resistance (AMR) is among the top 10 worldwide public health issues. AMR globally remains a public health concern, including in developing countries. (8) Like other countries, especially Zanzibar, Tanzania faces a critical challenge to AMR. (9–11) This fact poses a significant problem in Zanzibar, particularly for public healthcare facilities. Inappropriate use of antibiotic prescribing is the main factor contributing to antibiotic resistance, treatment failure, reduced antibiotic efficacy, rising healthcare costs, an increase in adverse medication, more extended hospital stays, and increased mortality rates (3,4). According to WHO estimates, half of patients do not take their medications as recommended, and over half of all medicines are improperly sold, administered, or prescribed. (3) Antibiotic medication abuse, overuse, or underused wastes limited resources and poses a public health risk. Globally, in 2019, 4.95 million people died from infections caused by infection; those 1.27 million casualties were directly related to AMR. In Tanzania in 2019, there were 54,000 deaths caused by AMR and 12,500 deaths directly related to AMR. Out of 204 nations, Tanzania has the 175th-highest age-standardized death rate per 100,000 people associated with AMR. (12) Inappropriate and irrational prescribing of antibiotics is a growing concern globally, leading to increased antimicrobial resistance, higher healthcare costs, and poor patient outcomes. In Zanzibar, Tanzania, limited data exists on the extent to which antibiotic prescribing in public healthcare facilities aligns with standard treatment guidelines and principles of rational drug use. It is difficult to implement effective interventions or policy changes without proper assessment. Several studies have assessed antibiotic prescribing practices in Tanzania; however, most of these studies have been conducted on the mainland. There is a lack of data, and no known study specifically evaluates antibiotic prescribing patterns in Zanzibar. This gap in research makes it difficult to understand the local prescribing practices, assess adherence to standard treatment guidelines, and implement targeted interventions.

Therefore, there is a critical need to evaluate the prescribing patterns of antibiotics in public health care settings in Zanzibar to identify gaps, promote rational use, and curb the rise of antimicrobial resistance.

Like other countries, Zanzibar mitigates this issue by improving public education on the responsible use of antibiotics, strengthening regulations to control antibiotic distribution, enhancing surveillance of antibiotic-resistant infections, and collaborating efforts with healthcare providers on rational use of antibiotics and prescribing (10). However, Zanzibar implements the sensible use of antibiotics. The information on the rational use of antibiotic prescribing is not well known, so this study will assess the rationale of prescribed antibiotic medicine in public healthcare facilities and identify any gaps/areas for improvement. The findings will help ensure that only antibiotics are provided when necessary. The current study aimed at achieving the following objectives;

1. To assess the prescribing practices of antibiotic medicine for prescribers in the Health facilities.
2. To determine the prescriber’s knowledge, perceptions, and attitudes on the rational antibiotic medicine prescribing in Health facilities
3. To find out key areas associated with the rational use of antibiotics and facilitate the prescription of antibiotic medicines in healthcare facilities, including barriers and challenges that contribute to irrational antibiotic prescribing, such as a lack of adherence to guidelines and antibiotic availability.

## 2. Materials and Methods

### 2.1 Research design and Approachj

A descriptive and cross-sectional study with a quantitative method approach was conducted retrospectively to assess the appropriate prescribed antibiotic medicines. A quantitative approach is defined as the collection of data that is suitable for numerical analysis. The cross-sectional design helps plan or administer control of the rational use of antibiotic prescribing. It can also help measure the prevalence/indicator of rational use of antibiotic prescribing, or a risk factor, and describe the features of a population. This method was achieved through the sensible use of quantitative approach. It was designed according to the objectives of the study.

### 2.2 Research location

The Zanzibar Island, a semi-autonomous area of the United Republic of Tanzania, served as the site of this study. It is situated in the Indian Ocean, 25 to 50 kilometres off the east coast of the Tanzanian mainland, between the longitudes of 39’19 79’.00 E and -6 ’16’ 39’.00 S. As stated by the 2022 Population and Housing Census, Zanzibar has a total area of 2461 km2 and a population of 1,889,773 people. Pemba and Unguja are the two largest islands in Zanzibar. (6)

The healthcare system in Zanzibar is divided into primary, secondary, and tertiary levels. One hundred nineteen primary health care units (PHCU) and 74 primary health care centers (PHCC) comprise the primary level, and 11 district hospitals and one regional hospital comprise the secondary level. The highest care delivery level is tertiary, with just one national referral hospital. As a result, Zanzibar has 206 public health facilities. (22)

The study was conducted at public health care facilities in the western urban region of Zanzibar from January 2023 to December 2023. The western urban region has thirty (30) public healthcare facilities owned by the Government through the Ministry of Health, including 8Health Centers and twenty-two (22 Dispensaries.

The study was conducted at public healthcare facilities in the western urban region of Zanzibar. This public health facility was selected because the western urban region has a problem of increasing_antimicrobial resistance due to the irrational use of antibiotic medicine. There is also a presence of more than one prescriber in the health facilities and relatively better availability of antibiotic medicines, hence creating a good situation for conducting an assessment of the rational use of antibiotic medicines prescribing.

### 2.3 Target and Study population

The target was all public healthcare facilities in the western urban region of Zanzibar, including thirty (30) health facilities, all prescribers, and all prescriptions prescribed during the study period. This study involved all prescriptions from the outpatient department from January 2023 to December 2023, and prescribers who prescribed in the outpatient departments were included. The eligible prescriptions and prescribers who fulfilled the inclusion and exclusion criteria were enrolled in the study.

### 2.4 Inclusion and exclusion criteria

#### Inclusion criteria

The Government owns all public healthcare facilities in the western urban region of Zanzibar through the Ministry of Health. Only OPD prescriptions from January 2023 to December 2023. The study included all registered prescribers working in public health facilities willing to give written informed consent. All prescriptions that prescribe antibiotic medicine.

#### Exclusion criteria

Emergency department prescriptions were not included in the study. New district hospitals, which were started in 2023, were not included in the study. Antibiotics for topical use were excluded from the study. All prescriptions with incomplete information were not included in the study.

### 2.5 Sample size

In compliance with WHO recommendations, at least 600 prescriptions should be used in a cross-sectional study describing the current prescribing practice of a health facility. (4,7,23) In this study, all prescriptions prescribed between January 2023 and December 2023 were calculated at the Outpatient department in selected thirty (30) public health facilities. Prescriptions were selected by using probability systematic sampling techniques from selected thirty (30) public health facilities.

K=N/n= the interval size, N= total number of prescriptions, n= sample size of the prescription. This formula was used to select the prescriptions. The sample size was expected to be 4320 prescriptions (120 prescriptions for each of 30 health facilities).

All the participants who enrolled in the study during this period filled out a structured questionnaire to measure the rational antibiotic prescribing and prescribing practice, including knowledge, attitude, and perception among prescribers towards the rational use of antibiotics. According to John W. Creswell, the minimum sample size was 15 participants in qualitative inquiry and research design. (24) Participants-prescribers were at least 15 participants. The participant-prescribers were selected by Convenience sampling.

### 2.6 Data collection instruments (Research instrument)

In this study, two instruments were used to collect data. One, to assess the prescribing practices of antibiotic medicine for prescribers in the health facility, and two, to determine the prescribers’ knowledge, perception, and attitude regarding the rational use of antibiotics.

Data was collected from patients’ medical records or prescription registration books. A structured data collection form adopted from the WHO/INRUD prescribing indicators guidelines and similar literature was used after modifying it to fit the current study. The first data collection instrument includes the following data:

The data include the number of patients, date, age, sex, prescribing frequency of antibiotics, dosage form, and duration. The data also include the percentage of antibiotics prescribed, the average number of medicines prescribed for each encounter, the proportion of prescriptions written under generic names, the percentage of prescriptions for antibiotics, the percentage of prescriptions for injections, and the percentage of prescriptions from the EML.

As shown in Appendix 1, a prescription data collection instrument was used to gather the required data from the January 2023 to December 2023 prescriptions.

Different questions were included in the questionnaire. The Kobo collect tool was used to collect data intended to evaluate the prescriber’s knowledge and attitude. It was focused on the prescribing practice and prescriber perception. Appendix 2

The data collection instruments were in electronic format, pre-tested, corrected, and printed. The researcher assistant has received training in data collection techniques, ethics, and research methodology.

### 2.7 Pre-testing

Two public health facilities were randomly selected through a computer used to pre-test in Microsoft Excel. This pilot study used a computer program to test, which involved a pre-test of prescription collection tools and a questionnaire on prescribing practice and prescriber perception. The data were then carefully evaluated to ensure they were accurate and comprehensive enough to meet the study’s objectives.

### 2.8 Validity

Validity relates to the degree to which a research instrument captures the intended data. This was done by applying identical procedures and operating the research instruments in a pilot study that followed the same criteria. Descriptive statistics were used to analyze the data and assess whether the study’s objectives were met.

Pre-testing was used to locate internal discrepancies in the research tool that could lead to measurement bias. Extensive training beforehand for the study instruments and data gathering method prevented this. Pre-testing the study tool helped to avoid measurement bias.

### 2.9 Reliability

Cronbach’s alpha, a metric used to assess the uniformity of research instrument outcomes, was used to characterize the data collection form’s internal consistency. The alpha value was found, confirming the study instrument’s reliability. Alpha range 0.7-0.9 (25)

### 2.10 Data collection procedure

Data was extracted from patients’ medical records or prescription registration books and questionnaires, assessing prescribing practice and determining prescribers’ perception of antibiotic prescribing at the selected public health facilities. The research assistant presented the study’s variables and summarized the data from the prescribing registration book and interview questionnaire at the public health facility. Structured questionnaires were used to collect the data during face-to-face interviews with interviewers. Data was collected using a Kobo Collect tool. The research assistant entered the data into the Kobo Collect tool. Kobo Collect data was exported to Microsoft Excel 2012 (MS Excel 10) for range and consistency checks, which were performed.

### 2.11 Data analysis

Clean data from MS Excel 10 was exported to the Statistical software package STATA v.15 for analysis. The descriptive statistics, including frequencies, percentages, averages (SD), and medians, were summarized using frequency distribution tables. Independent variables, such as demographic characteristics and outcome variables, were determined using chi-square. All survey responses were coded and analyzed using STATA v.15.

To meet the objectives, the analysis was divided into two main sections. The first section assessed the rational antibiotic prescribing and prescription practice questions adopted through the WHO prescribing indicator, and the second section included questions about prescribers’ knowledge, attitudes, and perceptions.

WHO/INRUD core indicators describing rational antibiotic prescribing were used, such as the average number of medicines prescribed for each encounter, the proportion of prescriptions written under generic names, the percentage of antibiotic prescriptions, the rate of injection prescriptions, and the percentage of prescriptions from the STG/EML. These indicators were calculated and summarized in tables, graphs, and charts.

### 2.12 Ethical Considerations

The Ministry of Health and Zanzibar Health Research Ethical Committee (ZAHREC) sought ethical approval to start the study. The Ministry was requested to grant permission to conduct the study. Some data was used retrospectively. However, to ensure their confidentiality, verbal consent was sought from the study participants before answering the questionnaire.

The questionnaire did not contain the participants’ names or personal information. Before analysis, the names and identifiers were removed from the dataset, which was stored and secured on my computer.

## 3. Results and Dscussion

This chapter presents and discusses the results of rational antibiotic medicine prescribing in public health facilities in Zanzibar, Tanzania, based on the research’s demographics and objectives. The data is presented in tables of percentages. A questionnaire was collected via the Kobo collect Tool, and data was collected from 70 prescribers and 3459 prescriptions in 30 public healthcare facilities.

### 3.1 Results

#### 3.1.1 Characteristics of prescribers

The study included 70 prescribers from 30 public healthcare facilities in the western urban region of Zanzibar. Most prescribers were clinical officers, 64 (91.4%), and the remaining were doctors, 6 (8.6%). Regarding gender distribution, 31.4% of prescribers were male, while 68.6% were female. The age of prescribers ranged from 21 to 57 years, with a median of 28 years. Regarding professional experience, years of practice among prescribers varied from 1 to 30 years, with a median of 4 years. Table 1 below provides a summary of these observations.

**Table 1:**
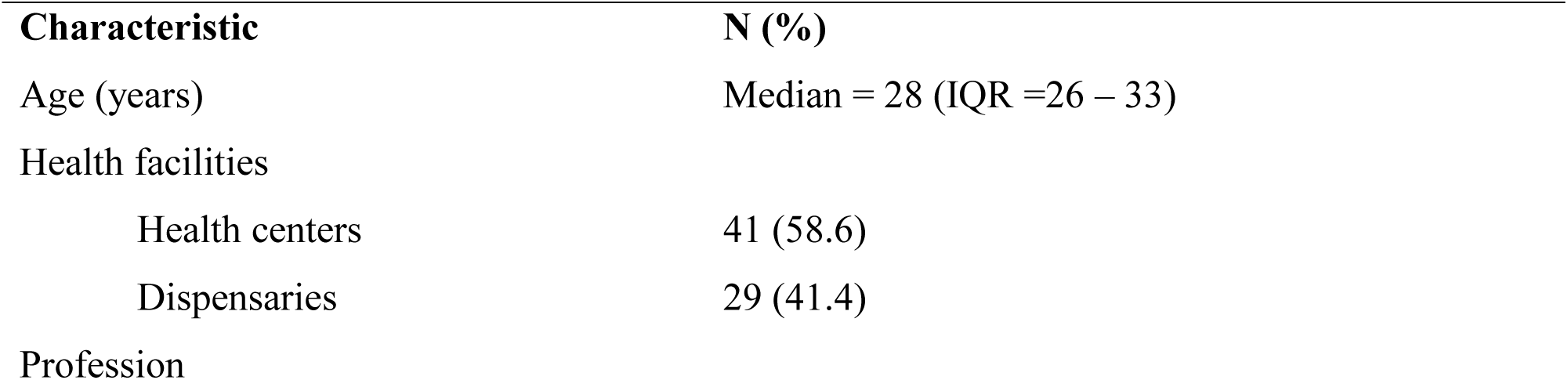
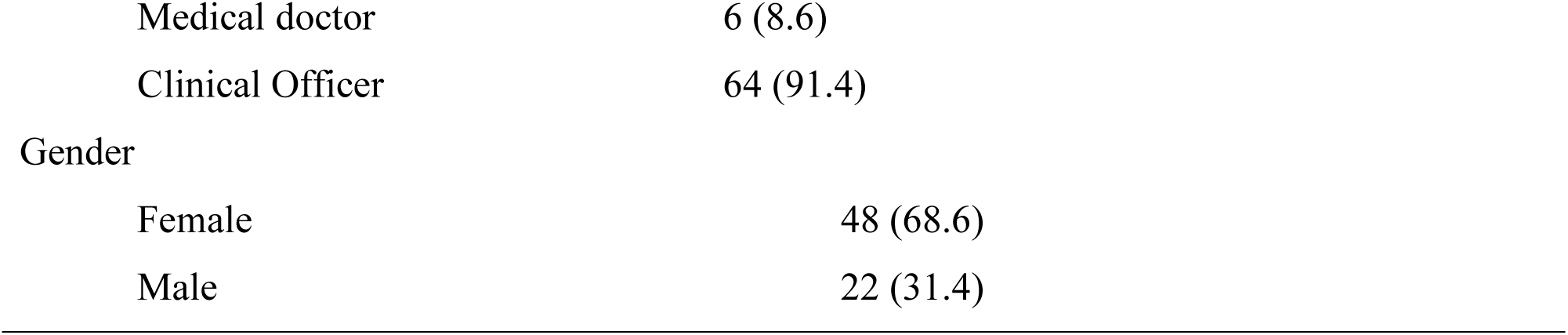
Prescribers’ characteristics.

| Characteristic | N (%) |
| --- | --- |
| Age (years) | Median = 28 (IQR =26 – 33) |
| Health facilities |  |
| Health centers | 41 (58.6) |
| Dispensaries | 29 (41.4) |
| Profession |  |
| Medical doctor | 6 (8.6) |
| Clinical Officer | 64 (91.4) |
| Gender |  |
| Female | 48 (68.6) |
| Male | 22 (31.4) |

Tables 2 show the distribution of prescribers’ sex in three districts. In West B and Urban, most prescribers are female, accounting for 68% and 62.9% respectively, while males constituted only 32% and 37% of the sample. Similarly, in West A, females represented 77.77% of prescribers, with males making up only 22.22%. This indicates that female prescribers are higher than male prescribers in the Health Facilities of these three districts.

**Table 2:** Gender distribution among the prescribers between the districts.

| DISTRICT | SEX |  | TOTAL |
| --- | --- | --- | --- |
|  | FEMALE | MALE |  |
| West A | 14 | 4 | 18 |
| West B | 17 | 8 | 25 |
| Urban | 17 | 10 | 27 |

Table 3 presents the educational characteristics of prescribers in three districts. In West A, West B, and Urban, most prescribers held diplomas (clinical officers), accounting for 94.4%, 84%, and 96.3% of the sample, respectively, followed by Bachelor’s degree holders at 5.55%, 16%, and 3.7%. In particular, no prescriber held a certificate or master’s degree in all districts. These results indicate that diploma holders are higher than Bachelor’s degree holders.

**Table 3:** Education characteristics among the prescribers between the districts.

| DISTRICT | PROFESSIONAL |  | TOTAL |
| --- | --- | --- | --- |
|  | CO | MD |  |
| West A | 17 | 1 | 18 |
| West B | 21 | 4 | 25 |
| Urban | 26 | 1 | 27 |

**Table 4:** Description of the assessed prescriptions.

| Characteristic | N (%) |
| --- | --- |
| District collected |  |
| West A | 831 (24.02) |
| West B | 1428 (41.28) |
| Urban | 1200 (34.69) |
| Sex |  |
| Female | 2163 (62.5) |
| Male | 1296 (37.5) |
| Patients' age (Mean) | 23.9 ±15.0 |

**Table 5:** Diagnosis underlying the use of Antibiotics.

| DIAGNOSIS | N (%) |
| --- | --- |
| URT | 883 (35.5) |
| UTI | 701 (20.3) |
| ENT | 598 (17.3) |
| Skin disease | 546 (15.8) |
| Diarrhea | 319 (9.2) |
| Dental disease | 119 (3.4) |
| Vaginal /Urethra discharge | 118 (3.5) |
| Cough/cold | 71 (2.1) |
| Conjunctivitis | 26 (0.8) |
| Other diagnoses | 42 (1.2) |
| Diagnosis not mentioned | 4 (0.1) |

**Table 6:** Most Utilized Antibiotics.

| S/N | ANTIBIOTIC | CLASS | PERCENTAGE (N%) |
| --- | --- | --- | --- |
| 1 | Amoxicillin | Access | 1185 (34.3) |
| 2 | Erythromycin | Access | 534 (15.4) |
| 3 | Ampicillin + Cloxacillin | Access | 523 (15.1) |
| 4 | Co-Trimoxazole | Access | 435 (12.6) |
| 5 | Metronidazole | Access | 221 (6.4) |
| 6 | Azithromycin | Watch | 199 (5.8) |
| 7 | Ceftriaxone | Watch | 199 (5.8) |
| 8 | Phenoxy methyl penicillin | Watch | 185 (5.3) |
| 9 | Doxycycline | Access | 107 (3.1) |
| 10 | Ciprofloxacin | Watch | 102 (2.9) |
| 11 | Cephalexin | Access | 30 (0.9) |
| 12 | Nitrofurantoin | Access | 22 (0.6) |
| 13 | Amoxiclav | Access | 14 (0.4) |
| 14 | Ampicillin | Access | 2 (0.1) |
| 15 | Amoxycillin + Flucloxacillin | Access | 2 (0.1) |
| 16 | Cefixime | Watch | 1 (0.0) |

#### 3.1.2 Characteristics of Outpatient Prescriptions

A total of 3459 prescriptions from outpatient departments across the 30 facilities were analyzed. The majority of these prescriptions were issued in West B (1,428; 41.3%), followed by Urban (1,200; 34.7%) and West A (831; 24%). The patient population had a mean age of 23.9 years (SD ±15.0), ranging from 1 to 96 years. The gender breakdown revealed that female patients constituted 62.5% (2,163), while males accounted for 37.5% (1,296).

Different diagnoses were documented, the most prevalent of which were Upper Respiratory Tract infections (URT), Urinary Tract Infections (UTI), skin-related conditions, and diarrhea as shown in 5 below.

Most Utilized Antibiotic

Different antibiotic medicines were prescribed, the most common of which were Amoxicillin, followed by Erythromycin and Ampicillin + Cloxacillin, as shown in 6 below.

Aware classification of Antibiotics.

Overall, 68.75% of antibiotics prescribed were in the Access class, with Watch class accounting for 31.25%, and no antibiotics were prescribed from the Reserve of antibiotics.

### 3.3 : Observed values of the prescribing practices indicators

3,455 (99.9%) recorded prescription encounters had a documented diagnosis, indicating comprehensive record-keeping. On average, 1.1 (SD=±0.3) antibiotics were prescribed per prescription encounter, with 2,277 (65.9%) prescribed generic names. Injectable antibiotics were recorded in 166 (4.8%) of prescriptions, while 2,270 (65.7%) of prescriptions adhered to the Zanzibar Standard Treatment Guidelines (STG). These findings are presented in Table 7.

**Table 7:** Observed values among the assessed prescriptions.

| Prescribing indicators | Observed values N (%) |
| --- | --- |
| Level of recording diagnosis | 3455 (99.9) |
| Average number of antibiotics per encounter | 1.1 |
| Prescriptions with more than one Antibiotic | 217 (6.27) |
| Antibiotics are prescribed with generic names | 2277 (65.9) |
| Encounters with injection-prescribed | 166 (4.8) |
| Antibiotics prescribed as per STG | 2270 (65.7) |

**Table 8.** Index of Rational Drug Prescribing (IRDP)

| Prescribing indicators | Observed values | WHO optimal value | Index for RDP (IRDP) |
| --- | --- | --- | --- |
| Level of recording diagnosis | 99.9% | 100% | 0.999 |
| Average number of antibiotics per encounter | 1.1 | <2 | 1.8 |
| Antibiotics are prescribed with generic names | 65.9% | 100% | 0.659 |
| Encounters with injection-prescribed | 4.8% | <25 | 1 |
| Antibiotics prescribed as per STG | 65.7% | 100% | 0.657 |

The calculation for the index for each indicator:

Injection and rational antibiotic and medicine per encounter, the following formula was applied: Index”WHO optimal value divided by observed value”.

For the generic name, EML/ STG, and diagnosis, the following formula was applied: Index “observed value divided by Optimal value”

### 3.4 : Knowledge, Attitude, and Perceptions of Prescribers

The prescribers’ knowledge of rational antibiotic prescribing was assessed using eight key questions from the data collection tool 1, with responses scored as 1 for correct answers and 0 for incorrect or neutral responses. The overall mean knowledge score among prescribers was 6.1 (SD = 0.89). 24.3% of prescribers had low knowledge (≤5 points), 41.4% had medium knowledge (6 points), and 34.3% had high knowledge (≥7 points). Table 9 should present the distribution of knowledge scores among prescribers.

**Table 9:** Knowledge scores distribution.

| Knowledge score range | Number of prescribers n (%) | Mean score (SD) |
| --- | --- | --- |
| Low knowledge (0 – 5) | 17 (24.3) | 4.88 (0.33) |
| Medium knowledge (6) | 29 (41.4) | 6 |
| High knowledge (7 and above) | 24 (34.3) | 7.13 (0.34) |

Prescribers’ attitudes toward rational antibiotic prescribing were assessed using a five-point Likert scale, ranging from 1 (Strongly Disagree) to 5 (Strongly Agree), with certain negatively worded statements reverse-coded to ensure consistency in interpretation. After adjusting for reverse-coded items, the overall mean attitude score was 7.5 (SD ±1.6).

The prescribers’ perceptions of rational antibiotic prescribing were assessed using yes/no questions. A majority of prescribers (90%) reported that they always adhere to the Zanzibar Standard Treatment Guidelines (ZSTG) and Zanzibar Essential Medicines List (ZEML) when prescribing antibiotics. 1.4% of prescribers acknowledged that they sometimes prescribe antibiotics even when unnecessary. Regarding antibiotic selection, 45.7% of prescribers prescribe only generic antibiotics. The study also found that beta-lactam antibiotics were the most prescribed, with 98.6% of prescribers confirming their frequent use. Table 10 summarizes the findings.

**Table 10:** Prescribers’ perceptions.

| Perception Statement | Yes (%) | No (%) |
| --- | --- | --- |
| Do you prescribe antibiotics when they are not necessary? | 1 (1.4) | 69 (98.6) |
| Do you prescribe only generic names of antibiotics? | 32 (45.7) | 38 (54.3) |
| Are beta-lactam antibiotics the most commonly prescribed? | 69 (98.6) | 1 (1.4) |
| Do you prescribe antibiotics even if the patient doesn't need them? | 0 (0) | 70 (100) |
| Do you use or follow ZSTG/ZEML when prescribing? | 63 (90) | 7 (10) |
| Can antibiotic resistance be controlled through education? | 70 (100) | 0 (0) |
| Do you need education and regular training on antimicrobial resistance? | 69 (98.6) | 1 (1.4) |

### 3.5: Key areas associated with rational antibiotic prescribing. Barriers to rational prescribing

Among the prescribers surveyed, 90.0% (n=63) reported adhering to standard treatment guidelines (STG/EML) when prescribing antibiotics, while 10.0% (n=7) did not follow these guidelines. Regarding generic names in prescriptions, 45.7% (n=32) of prescribers reported prescribing only generic medicines, whereas 54.3% (n=38) did not. Most prescribers, 98.6% (n=69), expressed the need for additional education and training on antimicrobial stewardship, while only 1.4% (n=1) indicated no need for further training.

### 3.2 Discussion

#### 3.2.1 Prescribing practices in public health facilities-Prescribing indicators

The demographic characteristics results show 3459 prescriptions across the 30 facilities. The majority were issued in West B (1,428; 41.3%), followed by Urban (1,200; 34.7%) and West A (831; 24%). There was a higher representation of female patients in all districts, particularly in Female patients, where more representation revealed that, on average, one antibiotic was prescribed per encounter, which is within the recommended WHO standard of ≤2 antibiotics per prescription (7,26). The most prevalent diagnoses were Upper Respiratory Tract infections (URT), Urinary Tract Infections (UTI), skin-related conditions, and diarrhea. The most commonly utilized antibiotic medicines were Amoxicillin, Erythromycin, and Ampicillin + Cloxacillin. The percentage of level recording diagnosis was 99.9%, closer to the WHO optimal value of 100%, with an index of 0.999, closer to the optimum value (1). The 99.9% level recording diagnosis in this study is higher than reported from Uganda (90.72%) (23). It is ideal for all prescriptions to include a diagnosis to assist pharmacists with integrating the disease and the medications.

The World Health Organization strongly advises all prescribers to prescribe medications using 100% generic names to reduce dispensing errors and promote better communication among healthcare providers. However, according to the current study, only 65.9% of prescriptions were written using generic names, despite WHO’s emphasis on generic prescribing to ensure affordability and accessibility of essential medicines (27). Even though most prescribers follow accepted prescribing procedures, some continue to use brand names in their prescriptions. The calculated percentage of prescriptions with the generic name was 65.9% %, which is lower than the WHO recommendation of 100%; with an index of 0.659, far from the optimum value (1), thus irrational (poor rational prescribing). The 65.9% of prescriptions with generic names in this study is less than what is reported from Uganda (90.21%), Tanzania (84.4%), and Ethiopia (91.5%) (23,28,29). The proportion of prescriptions adhering to Zanzibar Standard Treatment Guidelines (STG) was suggestive of potential gaps in adherence that require targeted interventions.

The research findings show that the percentage of antibiotic prescriptions by the hospital formulary/STG was 65.7%. However, the study from Asmara, Eritrea, shows that all medicines (98.39%) were from the Eritrean EML and hospital formulary, and antibiotics were prescribed by 53%. (4) Other studies from Ethiopia show that the percentages of antibiotics prescribed from the EML of Ethiopia and using generic names were 100% and 97.6%, respectively. (3,7). The study from Tanzania shows that 46.4% of patients who had a prescription for medicine also had a prescription for at least one antibiotic. Furthermore, 92.2% of the antibiotics prescribed at the health facility level complied with the most recent Tanzanian standard treatment guidelines. (14). The calculated percentage of medicines found in the essential medicines/STG was 65.7%, which is lower than the WHO recommendation of 100%; with an index of 0.657, which is far away from the optimum value (1), thus irrational (poor rational prescribing). The 65.7% of prescriptions found in STG in this study is less than what is reported from Uganda (78.96%), Tanzania (97.6%), and Ethiopia (98.7%) (23,28,29). These studies were consistent with a similar survey conducted in Nigeria. The strength of these studies was compliance with WHO standards for the rational use of antibiotic prescribing. (2)

Beta-lactam antibiotics were the most commonly prescribed, with most prescribers confirming frequent use. This trend is consistent with findings from other low-resource settings where beta-lactams are frequently used due to their broad-spectrum activity and availability (27). However, increased reliance on specific antibiotic classes raises concerns about selective resistance pressure, possibly contributing to rising AMR rates if not properly managed (30). A study from Ethiopia and Eritrea found that penicillin was the most commonly prescribed antibiotic (38.5%). (7) However, compared with other studies, Canada’s report was different, showing that fluoroquinolones were the most frequently prescribed antibiotic class. (5). In this study, the most commonly utilized antibiotic medicines were amoxicillin, consistent with a similar survey conducted in Tanzania (28). There could be several causes for the high percentage of antibiotic prescriptions.

In this study, the percentage of injections prescribed was 4.8% and fell under the WHO optimal value of <25%, with an index of 1. The 4.8% of injections prescribed in this study is higher than what is reported from Tanzania (3.2%) (28) but lower than reported from Uganda (5.43%), and Ethiopia (7.2%)(23,29). It is best to avoid prescribing injections irrationally because they are more expensive than other dose forms, and administering them might be hazardous to one’s health. Because of their quick beginning of action, injections are crucial formulations in some essential situations, such as emergencies, when other options are impractical or cannot be absorbed by extravascular routes.

##### Prescribers’ Knowledge, Attitudes, and Perceptions on Rational Antibiotic Use

There are 70 prescribers from 30 public healthcare facilities in the urban west region of Zanzibar. Most prescribers hold diplomas, 64 (91.4%) clinical officers, and the remaining hold bachelor’s degrees, i.e., doctors 6 (8.6%). In terms of gender distribution, female prescribers 68.6% are higher than male prescribers 31.4%. The median age of prescribers is 28 years, and the median professional experience is 4 years.

The findings indicate that while most prescribers demonstrated moderate to high knowledge of rational antibiotic use, some had low knowledge levels. These findings are concerning as inadequate knowledge among prescribers has been strongly linked to inappropriate antibiotic use and an increase in AMR. Similar studies in sub-Saharan Africa have reported comparable knowledge gaps among healthcare workers, emphasizing the need for continued professional training and education programs to improve prescribing practices (31). The study was conducted in Tanzania. It showed enough knowledge among prescribers and dispensers at 81.5% and 79.6%, with positive attitudes at 31.5% and 81.5%, and poor practices at 70.4% and 48%. (21) Another survey from Tanzania shows that the participants from Kilosa (84.5%), Ilala (83.6%), and Kibaha (73.7%) were likely to strongly agree that people should only use antibiotics when prescribed by medical professionals. (19). Also, 94.4% of Delhi, India, shows that prescribers were likely to strongly agree that antibiotic resistance is a significant problem in their setting. (16)

Furthermore, prescribers’ attitudes toward rational prescribing were generally positive, with a high mean attitude score. However, few prescribers acknowledged prescribing antibiotics even when unnecessary, indicating that non-clinical factors, such as patient demand, may sometimes influence decision-making. This aligns with previous research highlighting that patient expectations and healthcare system constraints contribute to irrational prescribing (32)

Knowledge, attitude, and perception of prescribers are crucial to enhancing the rational use of antibiotics. A study from a hospital in Delhi, India, shows that over 80% of the participants strongly agreed that the distribution of antibiotics without a prescription or the sale of antibiotics over the counter should be regulated. Approximately 69.6% of the participants thought that self-medication and antibiotic misuse are the two leading causes of ABR. (16)

##### Key barriers to rational prescribing

Several barriers to rational prescribing were identified, including guideline adherence, patient pressure, and antibiotic availability. While most prescribers reported adhering to STG/EML, the remaining 10% admitted not following guidelines, indicating room for improvement. Limited accessibility to standard treatment guidelines and uncertainty in clinical decision-making have been reported in other studies as key reasons for poor adherence to STG (33)

Moreover, almost half of prescribers stated that they prescribe only generic names, while the remaining 54.3% did not. This could be influenced by patient familiarity with brand names or supply chain constraints. WHO recommends consistent generic prescribing to enhance drug accessibility and cost-effectiveness (34)

A crucial finding was that most prescribers expressed the need for additional education and training on antimicrobial stewardship. This aligns with studies showing that continuous medical education (CME) programs are critical in strengthening prescribers’ competencies and improving adherence to best practices (35). In this study, 98.6% of the prescribers need training on antimicrobial stewardship. In contrast, a study from South Africa shows that 65% of physicians need training on antimicrobial stewardship. (36)

Also, the research from Tanzania shows that the participants appreciated some healthcare professionals for their increased understanding and for addressing misconduct. They asked for additional training on antibiotic resistance, ongoing education, and external seminars. (15)

This study was liable to some limitations. First, self-reported data on prescribing practices may be subject to social desirability bias, where prescribers might overreport adherence to guidelines. This was mitigated by triangulating data with prescription audits to validate responses. Second, the study was conducted in public health facilities only, which may limit the generalizability of findings to private-sector prescribers. However, including multiple healthcare levels (health centers and dispensaries) enhances the representativeness of the findings. Lastly, the study relied on cross-sectional data, which captures prescribing practices at a single time point and does not assess changes over time. This limitation is acknowledged, and future studies should incorporate longitudinal designs to track prescribing trends and the impact of interventions.

## 4. Conclusion

Prescribers in public health facilities in Zanzibar demonstrate moderate knowledge of rational antibiotic use, but gaps persist in guideline adherence, generic prescribing, and antimicrobial stewardship training. Prescribing practices indicate an appropriate average number of antibiotics per prescription but highlight a need for improved adherence to STG and increased use of generic names. It is recommended that prescribers use the generic names of the medications and base their antibiotic prescriptions on the STG. According to the results, most of the prescribing indicators in these health facilities did not meet the WHO’s standard requirements. Only the average number of medicines per encounter was suitable (rational). Barriers such as patient pressure, supply chain constraints, and lack of regular antimicrobial resistance training remain key challenges to rational prescribing.

The study’s findings suggest that antibiotic resistance should guide our plans for a more reliable approach to inform and educate our prescribers about antimicrobial resistance.The results are crucial for informing antimicrobial stewardship with pertinent interventions to improve antibiotic prescribing practices and alter prescriber behavior.

### 4.1. The study Implications

Based on the findings,

CME programs on antimicrobial stewardship should be prioritized to address knowledge gaps. Establishment of programs for awareness-raising and training: Prescribers working in health facilities should receive ongoing education and workshops on the rational use of medications, particularly antibiotics. Ensure updated guidelines are easily accessible to improve STG adherence. Every prescriber’s station should have standard treatment guidelines and promote adherence.

Strengthening prescription practices through mandatory generic prescribing and audit mechanisms can enhance compliance. Public awareness campaigns are needed to reduce patient-driven antibiotic demand, and supply chain management should be optimized to prevent stock-outs. Improve documentation and record-keeping in health facilities to facilitate quality data collection.

The Ministry of Health should ensure that health centers have a digital system for recording patients’ histories.Interventions and techniques, including clinical decision-support systems, antimicrobial stewardship programs, training programs for medical professionals, and updated guidelines, aim to promote the rational prescribing of antibiotics.

### 4.2 Limitations and direction for the future studies

The current study focuses on the rational prescriptions of antibiotics in the western unguja region in Zanzibar using cross-sectional descriptive design. Future research should assess the long-term impact of training programs, private-sector prescribing behaviours, and interventions to curb unnecessary antibiotic use. Furthermore, future studies can expand the scale of the study and include more geographical area of zanzibar and use longtudinal study design and see whether the findings could be the same as the design used in this study. The creation of baseline information on Zanzibar’s antibiotic prescribing practices that can be utilized for upcoming studies or to assess the results of any changes that are implemented.

## Data Availability

The researcher obtained ethical clearance to conduct this study and the certificate is attached

